# AI-Enabled Privacy-Preserving Cardiac Diagnostics via Electrocardiograms

**DOI:** 10.64898/2026.01.28.26345049

**Authors:** Fairuz Shadmani Shishir, Christopher J. Harvey, Amulya Gupta, Amit Noheria, Sumaiya Shomaji

## Abstract

Electrocardiogram (ECG) is a widely available, non-invasive diagnostic tool used for cardiovascular screening and provides essential insights into heart rhythm, structure, and function. However, the high dimensionality of ECGs and their entanglement with demographic information pose challenges for building fair and privacy-preserving machine learning models. ECG signals inherently encode soft biometric attributes such as sex, age, and race, which may introduce bias and raise privacy concerns in data-sharing environments. To address these challenges, we propose a deep learning framework that learns clinically relevant ECG representations while suppressing sensitive demographic information. We leverage a variational autoencoder (VAE) with a dual-discriminator architecture. One adversarial branch reduces soft biometric encoding, while the other preserves clinically important discrimination of reduced left ventricular ejection fraction (LVEF). The privacy-preserving reconstructed ECGs reduced identifiability of soft biometrics by independent CNN models with AUROC for sex 0.59 (from original 0.79), age 0.63 (from 0.78), and race 0.57 (from 0.69), while retaining clinically-useful predictions like reduced LVEF 0.82 (from 0.86), left ventricular hypertrophy 0.72 (from 0.75), and 5-year mortality 0.67 (from 0.66). These findings demonstrate the effectiveness of our approach for retaining ECG data yet protecting patient privacy.

## Introduction

Cardiovascular disease (CVD) remains the leading cause of mortality worldwide, accounting for approximately 19.8 million deaths annually, representing 32% of all global deaths according to the World Health Organization^1^. Early and accurate diagnosis of cardiac conditions is essential for effective treatment planning and improved patient outcomes. Among the most widely used non-invasive tools in cardiac diagnostics is the electrocardiogram (ECG), which records the heart’s electrical activity and provides valuable insights into arrhythmias, myocardial ischemia, conduction abnormalities, and other aspects of cardiac structure and function. ECGs are fast, cost-effective, and accessible across a wide range of clinical settings, making them indispensable in both acute and chronic cardiovascular care. Furthermore, ECG derived features have increasingly been explored in computational models for automated detection and prognostic assessments of cardiac dysfunction, including reduced left ventricular ejection fraction (LVEF) and heart failure risk^2,3^. However, with the increasing evaluation of ECG data for purposes beyond clinical diagnosis, such as biometric authentication^4^ and medical data-sharing initiatives^5^, there has been a growing emphasis on addressing privacy and data security concerns^6^. Recent advancements in deep learning have shown that personal attributes, including biological sex and age, can be accurately inferred from ECG signals^7^. This highlights a significant vulnerability: even anonymized ECG data can inadvertently disclose identifying information, potentially compromising patient confidentiality.

Conventional anonymization techniques such as data masking or removal of explicit identifiers often prove insufficient, as demographic attributes may remain inferrable from the signal data itself^8^. This paper by Kaissis et al.^9^ argues that existing anonymization strategies often focus on removing metadata, may be insufficient in the face of increasingly sophisticated AI models capable of extracting identifying features directly from the raw modality ECG signals. Evidence of this threat has already emerged. For instance, Packhäuser et al.^10^ demonstrated that deep learning models could achieve re-identification rates as high as 95% on chest X-ray datasets, despite standard anonymization. Similarly, Schwarz et al.^11^ successfully reconstructed recognizable facial images from cranial MRI data with 83% accuracy. In the context of ECGs, Ghazarian et al.^12^ reported that a convolutional neural network (CNN) trained on ECG waveforms could re-identify patients with up to 99.7% accuracy in certain subgroups by mapping waveforms to individual identities. Collectively, these findings underscore the need to move beyond conventional anonymization paradigms, particularly in the context of data sharing in AI workflows. An ideal anonymized medical dataset should retain predictive value for clinically relevant outcomes while being futile for inferring demographic details such as age, sex, and race.

### Existing Research in Cardiac Signal Analysis

Traditional methods for ECG signal interpretation typically rely on handcrafted feature extraction and rule-based analysis^13^. These methods often use parameters such as heart rate variability, QRS duration, QT interval, or ST–T wave morphology to detect abnormalities^14^. These approaches suffer from several limitations: they require specialized expertise for interpretation, exhibit inter-observer variability, and often lack accuracy^15^. Recent advances in deep learning have demonstrated promising results in automated cardiac signal analysis, with convolutional neural networks (CNNs) achieving impressive performance in tasks such as segmentation and classification. The growing recognition of privacy vulnerabilities has catalyzed the development of de-identification techniques along two primary trajectories: synthesis-based approaches and structure-preserving methods. Synthesis methods generate artificial ECG signals maintaining clinical utility while eliminating identifying characteristics. Generative Adversarial Networks (GANs) have gained prominence, with Thambawita et al.^16^ pioneering adversarial networks for ECG generation. Recent work by Alcaraz and Strodthoff^17^ demonstrated superior performance using Structured State Space Models for high-fidelity signal generation. Despite these advances, synthesis methods face persistent challenges, including ensuring signal diversity and validating clinical relevance. The evaluation of privacy-utility trade-offs remains inadequately addressed, with limited standardized metrics for assessing both privacy protection and clinical utility preservation.

Structure-preserving de-identification offers an alternative paradigm that retains authentic physiological patterns while effectively removing identifying characteristics. Despite its potential, research in this domain remains relatively limited. Notable contributions include the work of Salar et al.^18^, who utilized Ordinary Differential Equation-based Generative Adversarial Networks (ODE-GANs) to generate structure-preserving electrocardiogram (ECG) signals, and Bennis et al.^19^, who proposed a K-nearest neighbor (KNN) based algorithm, Chronos, for maintaining temporal structure in de-identified biomedical data. Critically, these methods often lack real physiological conditions with artificial data and provide limited analysis of downstream task performance. Chen et al.^20^ demonstrated that standard deep learning models trained on medical signals inadvertently encode demographic information that can be extracted by adversarial models, raising significant privacy concerns. These findings highlight the urgent need for privacy-preserving approaches in cardiac signal analysis. Table 1 presents a comparative summary of existing techniques for privacy protection in ECG data.

**Table 1.** Comparison of Privacy Preserving Techniques in ECG Data.

| Technique | Objective | Advantage | Limitations |
| --- | --- | --- | --- |
| ECG Synthesis<sup>21</sup> | Generate realistic DeepFake ECGs with GANs | Mimics real ECGs without identity link | Privacy is not considered |
| ECG Synthesis<sup>17</sup> | Generate high-quality synthetic ECGs for privacy | Long signal generation | It does not fully replace real data |
| Adversarial Training<sup>18</sup> | Preserve ECG privacy while keeping structure | Maintains clinical utility | Not ideal for memory-constrained environment |
| ECG Anonymization<sup>19</sup> | Share ECG data while retaining statistics | Better performance and anonymization | Limited customization and testing |
| Adversarial Training<sup>22</sup> | Sex de-identified ECG data | Sex anonymization | Limited clinical prediction |

In recent years, Variational autoencoders (VAEs), first introduced by Kingma and Welling^23^, have emerged as a powerful framework in generative modeling, combining the strengths of autoencoders with probabilistic inference. The VAE architecture consists of an encoder network that maps input data to a distribution in latent space and a decoder network that reconstructs the input from samples drawn from this distribution. In the context of ECG signal analysis, VAEs offer particular advantages due to their ability to handle the temporal, multi-channel, and physiologically constrained nature of cardiac signals. The probabilistic latent space representation naturally accommodates the inherent variability in ECG morphology across different individuals while preserving clinically relevant features^24^. Leur et al.^25^ utilized VAEs to improve the explainability of deep neural network-based ECG interpretation by learning underlying factors of variation in ECG morphology (the FactorECG). Their approach aimed to enhance interpretability by providing a more transparent representation of ECG features. Harvey et al.^24^ performed a comprehensive evaluation of VAE variants for dimensionality reduction, extracting 30 latent variables from representative ECG beats and applying them to downstream cardiac classification tasks. These approaches underscore VAEs’ potential to extract meaningful representations from ECG data that correspond to clinically relevant cardiac properties while accounting for the inherent variability in physiological signals across populations. The ability to model both the mean and variance of latent distributions makes VAEs particularly well-suited for handling the natural variation in cardiac rhythms and morphologies observed across different demographic groups and pathological conditions. These properties and established precedents make VAEs particularly suitable for our dual objective of demographic attribute removal and cardiac health prediction, as they can learn to disentangle clinically relevant cardiac features from potentially biasing demographic characteristics while maintaining the probabilistic nature essential for robust medical decision-making.

### Our Contribution

In this paper, we present Privacy-Preserving VAE (PP-VAE), which simultaneously addresses the challenges of privacy preservation and accurate cardiac function assessment. Our contributions are multifaceted:

1. We introduce a VAE architecture, PP-VAE, that encodes and reconstructs from the latent representations to remove demographic identifiers (age, sex, and race) from cardiac signals while preserving clinically relevant predictions (reduced LVEF, LVH, 5-year mortality). Results show strong predictive performance with AUROC of 0.82 for reduced LVEF, 0.72 for LVH, and 0.67 for 5-year mortality, while reducing sex identifiability from 0.79 to 0.59, age from 0.78 to 63, and race from 0.69 to 0.57.
2. To the best of our knowledge, this is the first study to employ adversarial training on ECG data for multi-demographic feature de-identification, proposing an objective function that optimizes both fairness and clinical accuracy through tailored loss formulations.

Our work represents a significant advancement in privacy-preserving medical signal analysis, with immediate applications in cardiology and broader implications for other medical domains where balancing diagnostic accuracy and patient privacy is crucial. The proposed methodology aligns with emerging ethical guidelines for responsible AI in healthcare^26^ and addresses regulatory requirements for protected health information^27^.

## Results

We conducted a series of experiments to evaluate the PP-VAE’s ability to suppress demographic information while preserving clinically relevant features. To this end, we used the 30-dimensional latent representations learned by different VAEs as input to XGBoost classifiers for a set of downstream prediction tasks, providing empirical evidence for the suitability of the 30-dimensional latent space shown in Table 2. The prediction targets included clinical outcomes such as reduced LVEF, LVH, and 5-year mortality as well as demographic variables, e.g., age, sex, and race. To compare, we evaluated four models: a baseline VAE, a VAE with adversarial sex loss, the proposed PP-VAE, and the proposed PP-VAE trained with an additional LVEF prediction objective (PP-VAE+LVEF). The results show that adversarial training enables VAEs to suppress sensitive demographic information partially. For example, the VAE with adversarial sex loss reduced predictive accuracy not only for sex, but also for age and race, although to a limited extent. In contrast, the PP-VAE, which incorporates penalties for both age and race, achieved a more substantial removal of demographic information from the encodings. Importantly, PP-VAE and PP-VAE+LVEF preserved the clinical utility of the representations. These models yielded improved performance on reduced LVEF and LVH prediction tasks while keeping demographic predictability close to chance levels. The additional LVEF prediction objective in PP-VAE+LVEF further strengthened performance on clinical outcomes without compromising privacy preservation.

**Table 2.** Prediction performance of XGBoost models using VAE and PP-VAE encodings with demographic variables (i.e., penalizing encodings of age, sex, and race). Lower values (↓) for sensitive attributes indicate better privacy preservation; higher values (↑) for clinical outcomes indicate better predictive utility

| Model | Sex ↓ | Age ($\leq 50$<br>vs. $>50$) ↓ | Race (White<br>vs. Non-White) ↓ | Reduced<br>LVEF ↑ | LVH ↑ | 5-year<br>Mortality ↑ |
| --- | --- | --- | --- | --- | --- | --- |
| Classical VAE | 0.75 | 0.70 | 0.62 | 0.82 | 0.72 | 0.66 |
| Penalize(sex) | 0.58 | 0.62 | 0.58 | 0.80 | 0.70 | 0.64 |
| PP-VAE | 0.57 | 0.60 | 0.56 | 0.80 | 0.70 | 0.65 |
| **PP-VAE+LVEF**<br>**(Proposed)** | 0.58 | 0.59 | 0.57 | 0.82 | 0.72 | 0.65 |

For an additional experiment. We then trained convolutional neural network (CNN) models to predict the same outcomes as Table 2 to see if the PP-VAEs were able to obscure the demographic information in the raw signals themselves while maintaining the morphological and clinical data within the signals. Table 3 evaluates the prediction performance of the CNN models, and it demonstrates that the PP-VAE+LVEF is able to remove the demographic information while maintaining the clinical use, even for reconstructed ECGs. This is a major step in privacy-preserving data sharing between health centers, as the modified ECGs can be sent over without breaking HIPAA compliance guidelines in an anonymized way^28^.

**Table 3.** Prediction performance using CNN models.

| Model | Sex ↓ | Age ($\leq 50$<br>vs. $>50$) ↓ | Race (White<br>vs. Non-White) ↓ | Reduced<br>LVEF ↑ | LVH ↑ | 5-year<br>Mortality ↑ |
| --- | --- | --- | --- | --- | --- | --- |
| Original ECG | 0.79 | 0.78 | 0.69 | 0.86 | 0.75 | 0.66 |
| Classical VAE | 0.76 | 0.73 | 0.66 | 0.83 | 0.73 | 0.69 |
| PP-VAE | 0.57 | 0.60 | 0.57 | 0.74 | 0.68 | 0.62 |
| **PP-VAE+LVEF**<br>**(Proposed)** | 0.59 | 0.63 | 0.57 | 0.82 | 0.72 | 0.67 |

We also measured the error between the raw signal and reconstructed signals. The reconstruction performance of the PP-VAE demonstrates robust signal recovery across all cardiac cycle components, with notable variations in performance characteristics that align with the inherent morphological properties of each waveform segment, showed in Figure 1. The QRS complex exhibits the strongest overall performance with an *R*^2^ value of 0.83 and Spearman correlation of 0.75, indicating excellent preservation of the high-amplitude, morphologically distinct ventricular depolarization patterns, despite registering the highest mean absolute error (MAE = 34.79 *µ*V) due to its greater dynamic range. The T-wave reconstruction achieves the highest correlation coefficient (*ρ* = 0.79) with moderate error levels (MAE = 20.24 *µ*V) and strong explained variance (*R*^2^ = 0.78), reflecting the model’s capacity to capture the smooth, predictable repolarization morphology. In contrast, P-wave reconstruction presents the most challenging scenario with the lowest *R*^2^ (0.54) and correlation (0.64) values, though maintaining the smallest absolute error (MAE = 14.75 *µ*V), which is consistent with the inherently variable and low-amplitude nature of atrial depolarization that presents greater difficulty for autoencoder architectures to reliably reconstruct. All correlations demonstrate statistical significance (*p <* 0.001), confirming the model’s consistent ability to preserve essential ECG morphological features across the complete cardiac cycle.

**Figure 1.**
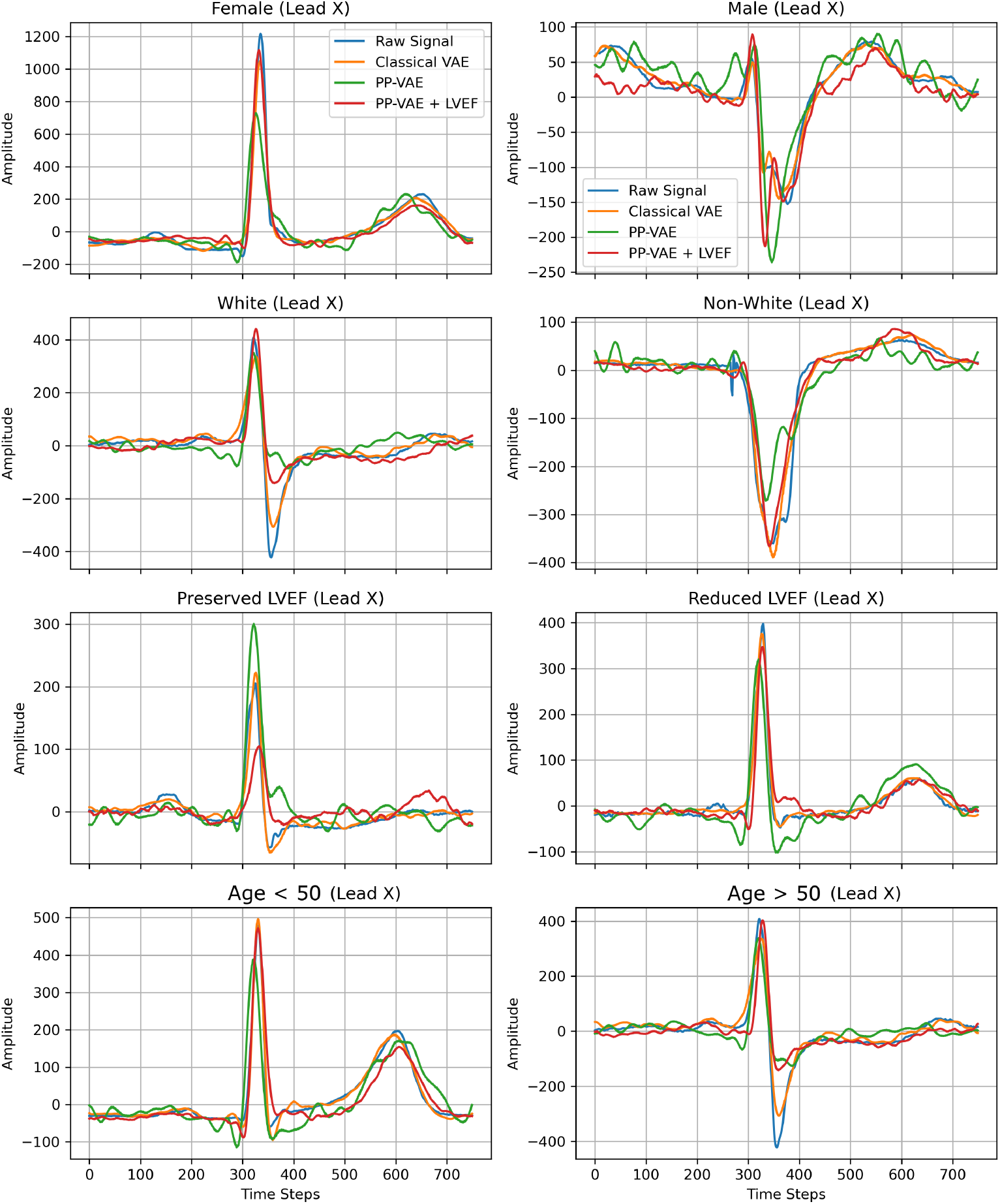
Representative Lead X signals are displayed for each subgroup, stratified by sex (female, male), race (white, non-white), left ventricular ejection fraction (LVEF; preserved, reduced), and age (under 50 years, over 50 years). Four signal types are shown: the raw ECG signal (blue), reconstruction from a classical Variational Autoencoder (VAE; orange), a physiologically-primed VAE (PP-VAE; green), and a PP-VAE conditioned on reduced LVEF (red). The figure demonstrates how different model architectures preserve signal morphology and amplitude across subgroups. Notably, the PP-VAE + reduced LVEF model yields improved reconstruction fidelity, particularly in cases with preserved LVEF and younger patients.

Figure 2 and Figure 3 illustrate the reconstructed median-beat ECG waveforms by sex and reduced LVEF category across different leads and model configurations.

**Figure 2.**
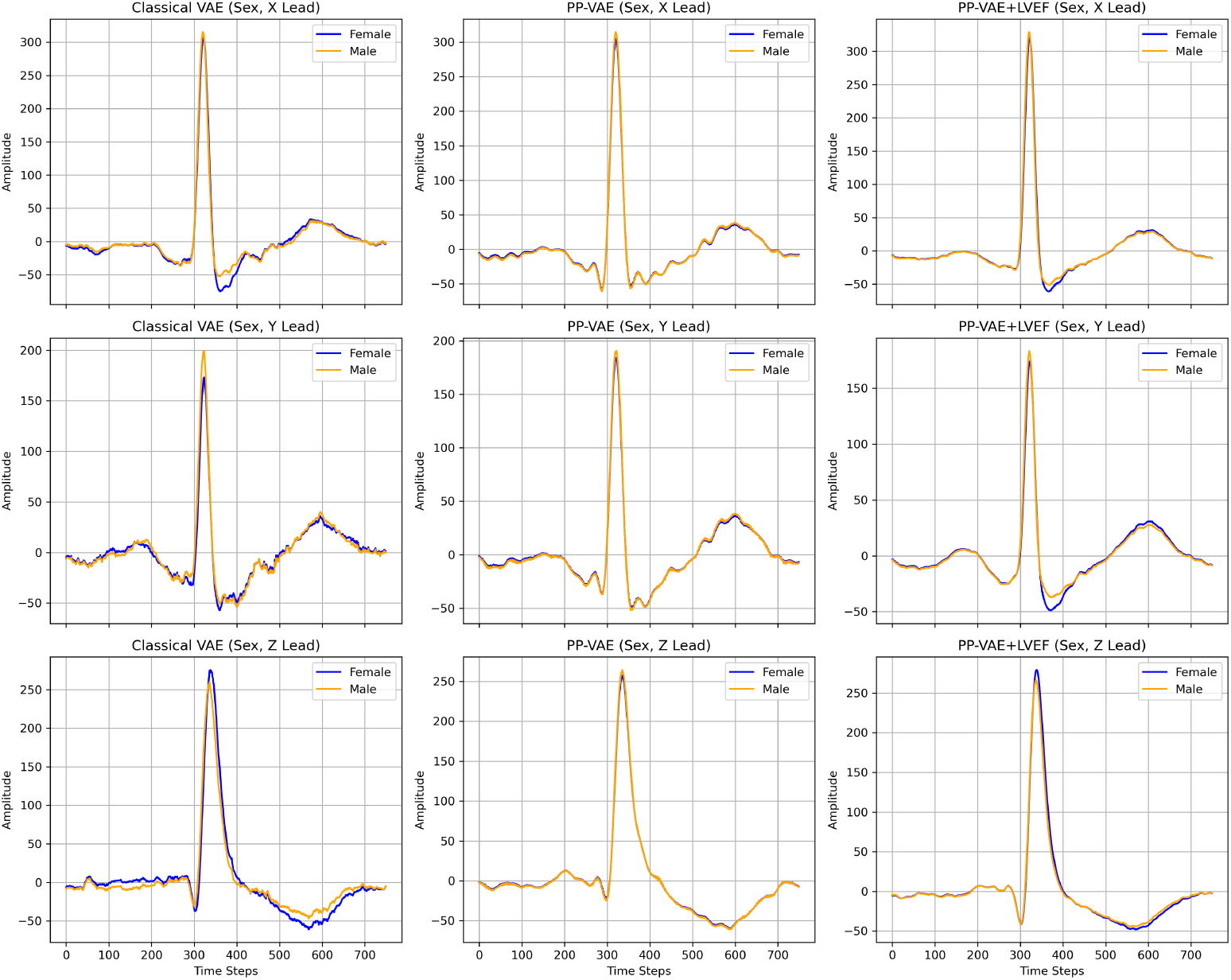
Reconstructed median-beat ECG waveforms by sex across different leads and model configurations. Each subplot shows the mean reconstructed ECG signal for female (blue) and male (orange) participants, plotted over time steps for three orthogonal ECG leads (X, Y, Z) using three model types: Classical VAE (left column), PP-VAE (middle column), and PP-VAE + LVEF (right column). The X-axis represents normalized time steps of the median beat, and the Y-axis represents the mean amplitude in *mVs*. Across all leads and models, the QRS complex appears as a sharp positive peak around time steps 320–350 *ms*, followed by characteristic T-wave morphology. Male and female waveforms are nearly overlapping in all cases, with minimal amplitude deviations, indicating that sex-specific differences in the reconstructed ECGs are largely attenuated by the models. PP-VAE and PP-VAE + LVEF yield smoother baselines compared to the Classical VAE, with slightly reduced post-QRS variability between sexes, particularly in the X and Z leads.

**Figure 3.**
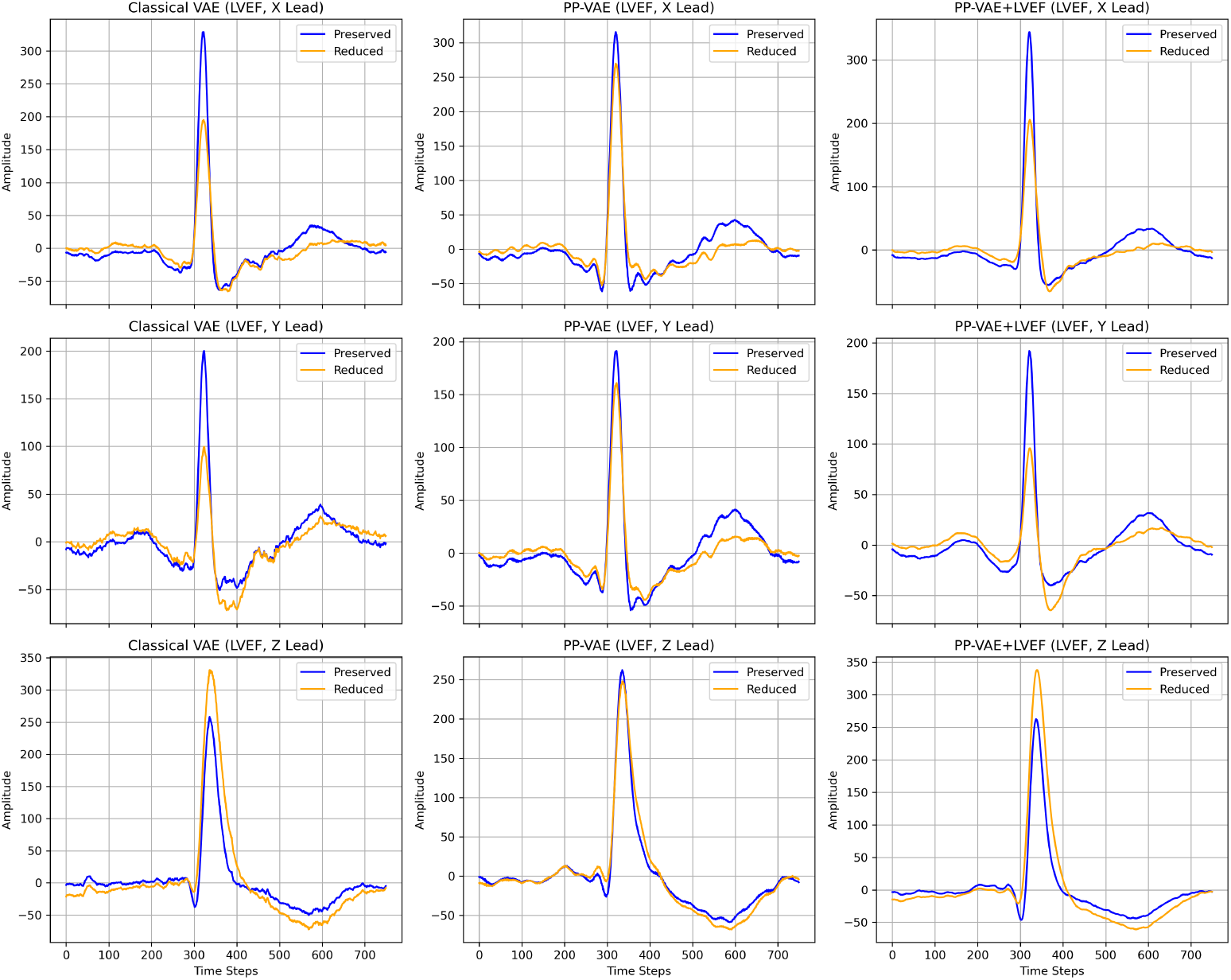
Reconstructed median-beat ECG waveforms by reduced LVEF category across different leads and model configurations. Each subplot displays the mean reconstructed ECG signal for participants with preserved LVEF (blue) and reduced LVEF (orange), across three orthogonal ECG leads (X, Y, Z) and three model types: Classical VAE (left column), PP-VAE (middle column), and PP-VAE + LVEF (right column). The X-axis represents normalized time steps of the median beat, and the Y-axis represents the mean amplitude in *mVs*. In all leads, the QRS complex appears as a sharp peak around time steps 320–350 *ms*, followed by the T-wave. Compared to preserved LVEF, reduced LVEF cases show lower T-wave amplitudes and, in some leads, subtle QRS morphology differences. These distinctions are most visible in the Classical VAE reconstructions, particularly in the X and Y leads, while PP-VAE and PP-VAE + LVEF tend to reduce between-group differences and produce smoother baselines.

## Discussion

In this work, we introduce a Privacy-Preserving Variational Autoencoder (PP-VAE) for ECG analysis. By integrating adversarial demographic obfuscation with clinically supervised representation learning, our framework produces latent embeddings that retain morphological and diagnostic information while suppressing sensitive demographic attributes. This dual objective enables the model to balance clinical utility with privacy preservation. While VAEs have demonstrated promise for dimensionality reduction, anomaly detection, and generative tasks in ECG analysis, existing implementations have largely been restricted to single-lead or standard 12-lead recordings in controlled environments. By contrast, our approach demonstrates feasibility in 10-second ECG recordings, highlighting its potential scalability. Nonetheless, most VAE-based methods, including ours, remain limited by their reliance on fixed-length segments, which constrains applicability to continuous monitoring where arrhythmic events may occur sporadically over extended periods.

Despite recent advances in interpretable latent representations^25,29^, the interpretability of VAE latent spaces continues to pose a barrier to clinical adoption. While certain dimensions can be aligned with canonical ECG features, the relationship between latent factors and specific pathophysiological processes often remains opaque. This interpretability gap constrains the capacity of such models to provide explanatory insights alongside predictions, a critical requirement for decision support in clinical settings. Another related technical challenge is the choice of latent priors. The conventional Gaussian prior in VAE may not adequately capture the complex manifold structure of ECG signals, potentially limiting both generative performance and representation quality. Although alternative priors have been proposed^30^, systematic evaluation is required to determine appropriate distributional assumptions for cardiac data. An additional consideration concerns the entanglement of clinical and demographic information. For example, age is strongly associated with reduced LVEF, meaning that ECG features predictive of age may also carry legitimate diagnostic value. Removing demographic correlates can therefore impair clinical prediction, not due to model failure but because of genuine biological associations. This highlights the importance of task-specific evaluation when attempting to mitigate demographic information. Moreover, the removal of demographic features does not inherently resolve issues of fairness; in some cases, it may exacerbate inequities. Differences in disease prevalence, phenotype expression, or comorbidity across demographic groups can alter conditional feature distributions, thereby harming subgroup performance or calibration. Careful fairness assessment, including subgroup error and calibration analyses, is therefore essential when applying privacy-preserving transformations. Further limitations arise from the lack of evaluation on conventional ECG summary statistics (e.g., QRS duration, PR interval, QTc). Understanding how these clinically interpretable measures are altered by transformation is crucial, as such changes affect both human interpretability and compatibility with rule-based diagnostic workflows. In the absence of correction or inverse mapping, transformed ECGs may become unintelligible to clinicians, presenting a significant challenge for deployment. Beyond interpretability, confidentiality and adversarial privacy risks also warrant consideration. Practical implementation requires strict protection of both the guiding classifiers and the original ECG data. Leakage of model weights, training data, or even aggregated statistics could enable adversaries to reconstruct demographic information. Furthermore, class imbalance in real-world datasets can implicitly encode demographic correlates, enabling partial re-identification. Addressing these vulnerabilities necessitates formal threat modeling and the integration of privacy-preserving techniques such as differential privacy and secure model hosting. Finally, the publication of de-identification methodologies presents a dual-use concern: while transparency promotes scientific progress, it may also enable adversaries with auxiliary datasets to replicate and invert de-identification pipelines. This underscores the need for complementary technical, procedural, and legal safeguards, rather than reliance on model-based transformations alone.

In summary, VAEs represent a powerful framework for ECG representation learning, yet their clinical translation depends on more than predictive accuracy. Progress in robustness to real-world artifacts, interpretability of latent spaces, and the integration of privacy- and fairness-preserving mechanisms will be critical for trustworthy adoption. Future work should focus on extending clinically supervised latent objectives to a broader range of outcomes, systematically evaluating the effect of transformations on standard ECG markers, and developing rigorous fairness and privacy assessments. With these complementary advances, representation-learning frameworks such as PP-VAE can move from experimental demonstrations to reliable components of precision cardiology pipelines.

### Future Directions

The present study highlights the potential of privacy-preserving VAEs for ECG representation learning, but also reveals several avenues for advancing both the methodology and its clinical applicability. Building on our findings, we identify the following promising directions:

- **Privacy-Preserving Learning:** Future work can integrate federated learning and differential privacy techniques to extend our framework beyond centralized datasets. This would enable multi-institutional collaboration while rigorously safeguarding sensitive patient data, a prerequisite for real-world deployment.
- **Physiologically Informed Priors:** Incorporating priors grounded in cardiac electrophysiology, such as distributions over RR intervals, QT durations, or QRS widths, has the potential to align the learned latent space with an interpretable, physiologically meaningful objective. This would improve model transparency and facilitate adoption by clinicians, who often demand correspondence between model features and known cardiac parameters.
- **Personalized Monitoring:** Current representations are optimized at the population level, yet clinical care often requires individualized assessment. Adaptive VAEs that establish patient-specific baselines could detect subtle deviations over time, particularly beneficial for monitoring individuals with chronic or high-risk cardiac conditions.
- **Fairness under Privacy:** While our work demonstrates that demographic attributes can be obfuscated, an open question remains: how do privacy-preserving constraints interact with fairness across demographic and clinical subgroups? Future efforts should design strategies to simultaneously mitigate bias in latent spaces and ensure equitable performance under privacy-preserving regimes.

## Conclusion

The results of this study demonstrate the potential of privacy-preserving deep learning for ECG analysis in clinical practice. Standard deep learning models achieve strong predictive performance for critical cardiovascular outcomes, including reduced LVEF, LVH, and 5-year mortality, yet inadvertently encode sensitive demographic information such as sex, age, and race, raising concerns about patient privacy and algorithmic fairness. To address this, we introduced Privacy-Preserving VAE and further extended it with task-relevant supervision (PP-VAE + LVEF). Our results demonstrate that the proposed model significantly reduces the encoding of sensitive attributes (e.g., Sex: 0.58 vs. 0.75 in classical VAE) while maintaining strong predictive performance on key clinical outcomes (e.g., LVEF: 0.82, matching classical VAE). This illustrates a favorable privacy-utility trade-off, showing that incorporating adversarial tasks into the VAE architecture and loss function can mitigate demographic leakage without sacrificing clinical relevance.

## Methods

### Dataset

We reduced each 12-lead 10-second ECG signal to a single 750-ms median representative beat, centered 100-ms after the QRS onset. Using the Kors regression matrix, we projected the 8 independent leads into orthogonal X, Y, and Z components. This transformation yielded a 2250-point signal vector sampled at 1000Hz. We normalized the signals to a -1 to 1 range using global absolute max scaling. A total of 1,065,368 ECGs were split by patient ID into 90% training and 10% holdout sets for model development and evaluation.

### Ethical and IRB Approval

All methods were carried out in accordance with relevant guidelines and regulations. Informed consent was obtained from all subjects and their legal guardians prior to participation. The study was approved as an expedited review by the Institutional Review Board at the University of Kansas Medical Center (STUDY00160252). It included all clinically acquired ECGs from the University of Kansas Health System between 2008 and 2022. When available, ECGs were linked to echocardiographic variables within 45 days of ECG using the Healthcare Enterprise Repository for Ontological Narration (HERON) via medical record numbers.

### Problem Formulation

We formulate the problem as a multi-objective optimization task: (1) learn a latent representation of cardiac signal that effectively predicts reduced LVEF, LVH, and 5-year mortality; (2) remove demographic attributes (age, sex, race) from the learned representation to ensure patient privacy; and (3) preserve sufficient morphological information for accurate reconstruction and interpretation. Let *x* represent the ECG input signals, *y*_*c*_ denote the clinical targets (reduced LVEF, LVH, and 5-year mortality parameters), and *y*_*d*_ represent the demographic attributes to be removed (age, sex, and race). Our goal is to create an encoder *E* that maps the input signals to a latent space *z* = *E*(*x*) such that *z* contains minimal information about *y*_*d*_ while maximizing information about *y*_*c*_.

### Privacy-Preserving VAE Architecture

Our model architecture, illustrated in Fig. 4, consists of four main components: (1) an encoder network, (2) a decoder network, (3) a clinical predictor network, and (4) demographic predictor network. The encoder network compresses the input signal data down to 30 latent data points, which are then fed to the decoder network to reconstruct. The weighted Mean Squared Error (w-MSE) between them is the reconstruction loss. The clinical and demographic predictor networks take in the 30 latent encodings from the input signal and use those to predict their respective targets. The loss from these predictor networks is then combined with the reconstruction loss to update the encoder and decoder networks. This design extends the traditional VAE framework to incorporate both privacy preservation and task-specific prediction.

**Figure 4.**
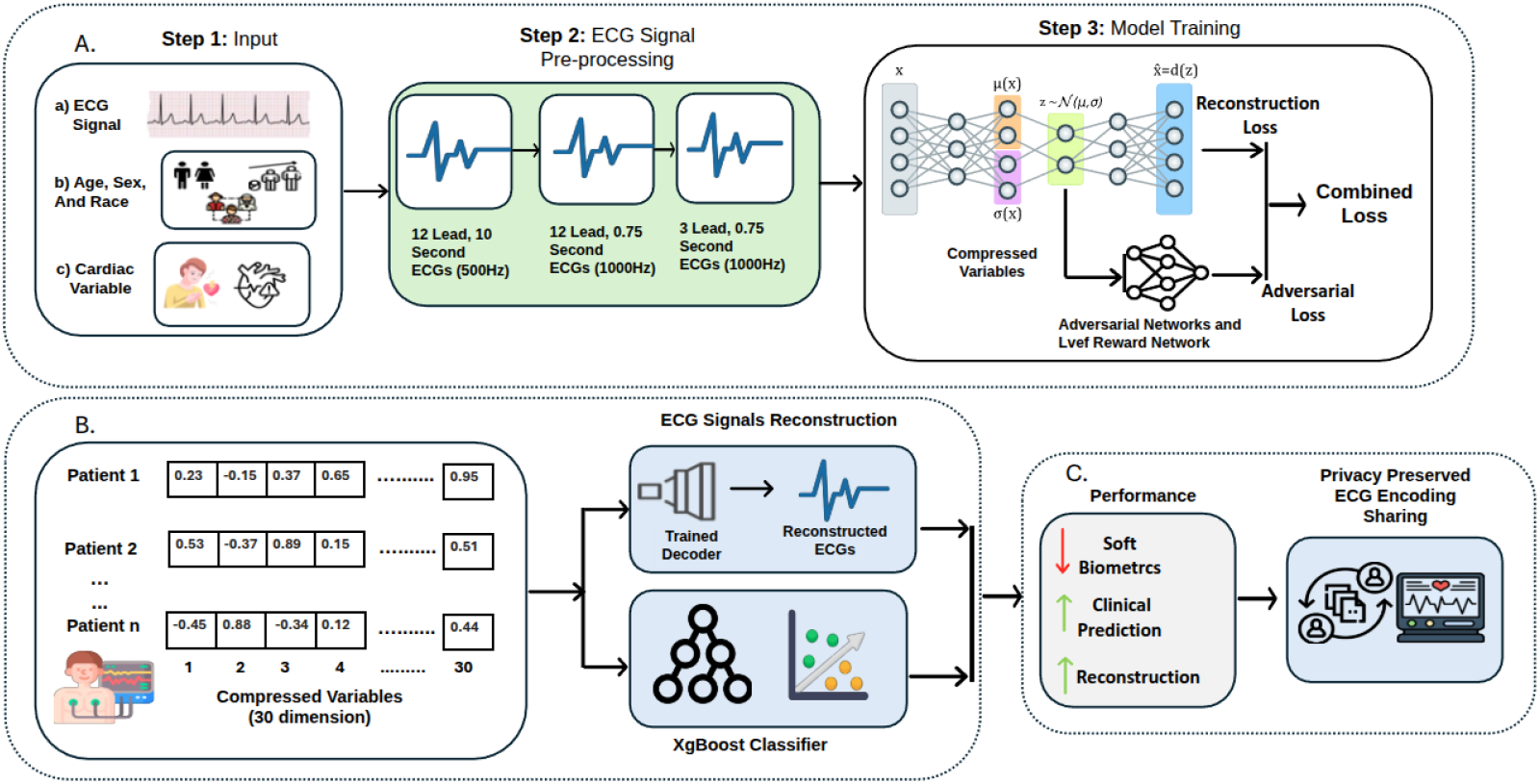
Overall architecture of the proposed framework. (A) Model development using VAE with adversarial training to remove demographic attributes from ECGs. (B) Performance analysis for downstream tasks. (C) Applications of the privacy-preserving ECG embeddings in clinical outcome prediction and secure data sharing.

### Encoder Network

The encoder maps input cardiac signals *x* ∈ ℝ to a latent distribution parameterized by mean *µ*_*z*_ and log-variance log 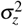.

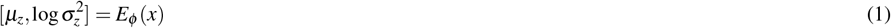

In the equation, *ϕ* represents the encoder parameters. The latent representation *z* is sampled using the reparameterization trick:

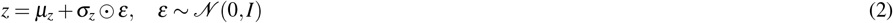

In the equation, 𝒩 (0, *I*) denotes the standard normal distribution with zero mean and identity covariance. The encoder is designed to process input tensors of shape (3, 750, 1) through a series of convolutional and fully-connected layers. It is made up of 4 2D convolutional layers with (32, 32, 64, 64) filters, a kernel size of 15, a stride of 2, tanh activations, and batch normalization. Follow by 2 fully-connected layers with (1024, 256) neurons, L2 regularization, tanh activation, and dropout of 0.25 between them. The last fully-connected layer is sampled to create the 30 latent variables, *z*.

### Decoder Network

The decoder reconstructs the input signal from the latent representation:

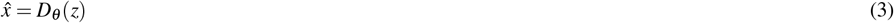

In the equation, *θ* represents the decoder parameters. The decoder reconstructs the input from the 30 latent variables. It is the inverse of the encoder network. It has 2 fully connected layers (256, 900) neurons with L2 regularization, tanh activation, and dropout of 0.25 between them. The output is reshaped into a (3, 300, 1) matrix and fed into 4 2D Convolutional Transpose layers with (128, 64, 32, 3) filters, kernel size of 15, strides of [(1,1), (1,1), (1,2), (1,2)], linear activations, and batch normalization. The final transposed convolutional layer produces the reconstructed input sequence.

### Clinical and Demographic Predictor Networks

The clinical and demographic predictor networks share the same architecture. The input to the networks is the 30 latent encodings. These are then fed to 4 fully connected layers with (256, 128, 64, 32) neurons, ReLU activation, and dropout between layers of 0.2. The final output layer is a single neuron with a sigmoid activation function to produce a probability score. Separate instances of this discriminator are trained with distinct optimizers to predict each attribute (sex, age, race, and reduced LVEF).

## Loss Function Description

The overall loss function integrates multiple objectives to optimize the latent representation for clinical relevance while minimizing demographic information. Below, we detail each component of the loss function.

### Reconstruction Loss

To better capture clinically relevant ECG morphology, we designed a w-MSE reconstruction loss. Instead of treating all parts of the signal equally, we give different importance to the P wave, QRS complex, and T wave, and the loss demonstrates:

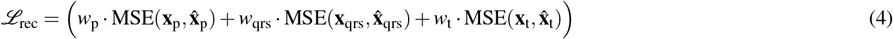

where *w*_p_, *w*_qrs_, *w*_t_ are manually set weights (20, 10, and 15, respectively). These values were chosen to counterbalance intrinsic amplitude differences across ECG components: the P wave (lowest amplitude) was given the highest weight, the QRS complex (highest amplitude) was assigned the lowest weight, and the T wave was weighted in between. This design ensures that low-amplitude but clinically significant features are not overshadowed during reconstruction, while still maintaining fidelity to the overall ECG morphology.

### Adversarial Losses for Demographic Attributes

Adversarial losses are employed to minimize the information about demographic attributes (gender, age, race) in the latent representation **z**. Each loss is computed using binary cross-entropy and scaled by a hyperparameter *λ*_adv_ which was set to 0.5.

### Gender Adversarial Loss

The gender adversarial loss aims to make the latent representation **z** uninformative about gender. The gender label is binarized as follows:

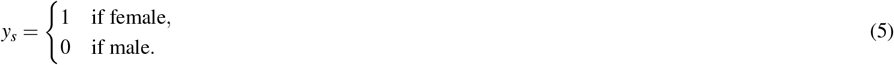

The adversarial loss is defined as:

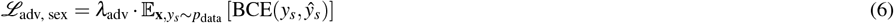

where:

- *λ*_adv_ is a weighting hyperparameter that controls the strength of the adversarial penalty relative to the other loss components.
- **x** denotes the input ECG signal, and *y*_*s*_∈ { 0, 1} represents the ground-truth sex label sampled from the empirical data distribution *p*_data_.
- **z** is the latent representation produced by the encoder.
- *ŷ*_*s*_ = *D*_sex_(**z**) is the predicted probability of sex from the adversarial discriminator *D*_sex_.
- BCE(·, ·) is the binary cross-entropy loss, which measures the discrepancy between the true label *y*_*s*_ and the discriminator’s prediction *ŷ*_*s*_.

### Age Adversarial Loss

The age adversarial loss processes the age label by converting it to a binary variable (e.g., indicating “older than 50” or “50 and below”):

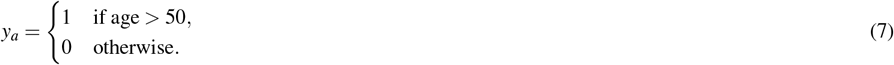

The loss is then:

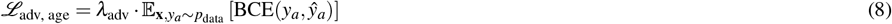

where *ŷ*_*a*_ = *D*_age_(**z**) is the predicted binary age label from the age adversarial discriminator.

### Race Adversarial Loss

The race adversarial loss processes the race label by converting it to a binary variable (e.g., indicating “white” or “non-white”):

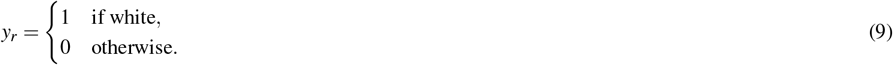

The loss is then:

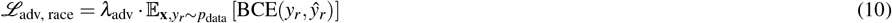

where *ŷ*_*r*_ = *D*_race_(**z**) is the predicted binary race from the race adversarial discriminator.

### Reduced LVEF Prediction Loss

The reduced left ventricular ejection fraction (reduced LVEF) prediction loss ensures that the latent representation **z** retains clinically relevant information. The LVEF value is binarized as follows, using a cutoff of 40% motivated by clinical convention^31^

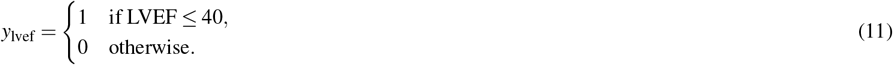

The prediction loss is defined as:

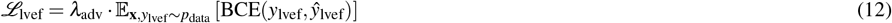

where *ŷ*_lvef_ = *P*_lvef_(**z**) is the predicted binarized LVEF from the predictor network.

### KL Divergence Loss

The KL divergence loss regularizes the latent space by encouraging the approximate posterior *q*(**z**| **x**) to be close to a standard normal prior *p*(**z**) = 𝒩 (0, *I*):

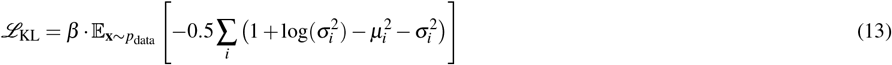

where *µ*_*i*_ and 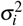 are the mean and variance of the latent variable **z**, and *β* is a hyperparameter controlling the strength of regularization. The *β* value for the PP-VAE+LVEF was 3 after empirical evaluation.

### Combined Loss Function

The total loss for the PP-VAE+LVEF model combines all components, balancing reconstruction, adversarial, clinical prediction, and regularization objectives:

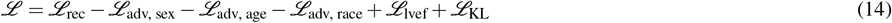

Our training objective is designed to explicitly separate clinically relevant representations from demographic information while preserving the morphological structure of the signal. To achieve this, we adopt an adversarial learning scheme. By subtracting the demographic adversarial loss from the overall objective, we effectively encourage the encoder to learn latent variables *z* from which demographic classifiers perform poorly. In other words, the optimization process forces *z* to be invariant to demographic attributes, thereby obfuscating sensitive information. At the same time, we include two positive objectives: (i) the left ventricular ejection fraction (LVEF) prediction loss, and (ii) the reconstruction loss. The prediction term ensures that *z* retains clinically meaningful information necessary for downstream diagnostic tasks, while the reconstruction term ensures that *z* captures morphological detail required to faithfully represent the input signal. These two forces together anchor the latent space to clinically relevant and morphologically faithful embeddings, preventing the model from discarding too much information in pursuit of privacy. This multi-objective formulation reflects our design choice: we subtract adversarial demographic losses to maximize obfuscation of sensitive attributes, while simultaneously adding prediction and reconstruction losses to preserve utility. As a result, the PP-VAE learns a latent representation that balances privacy preservation (via demographic invariance) with clinical utility (via LVEF prediction) and signal fidelity (via reconstruction).

### Training Strategy

We first pretrained the encoder and decoder networks in a normal VAE on 1 million ECGs. This helped make the model more robust and already understand what ECG encodings should look like. We then applied these weights to the PP-VAE’s encoder and decoder and trained for an additional 200 epochs. The encoder, decoder, and predictor networks were all trained simultaneously, each epoch on 4 NVIDIA V100 GPUs. The model was implemented using TensorFlow. We used the Adam optimizer with a learning rate 10^−6^ and a batch size was 32. The latent space dimension was set to 30 based on ablation studies measuring reconstruction quality and prediction accuracy.

### Validation Using State-of-the-Art Model

Following the training of our PP-VAE, we validated the encoded outputs using a slightly modified version of the state-of-the-art CNN model architecture proposed by Attia et al.^7^, developed at Mayo Clinic on 280,000 raw ECG signals. Table 3 summarizes the performance of this model in predicting both demographic and clinical attributes from the original and VAE-reconstructed ECG signals. As expected, the original ECG signals yielded strong predictive performance on the raw signals, especially for reduced LVEF (AUC = 0.86) and age estimation (AUC = 0.78). When tested on reconstructed signals from a classical VAE, the model exhibited a moderate decline in performance, though the degradation was minimal.

Importantly, our PP-VAE markedly suppressed the CNN’s ability to infer sensitive demographic attributes—such as sex (AUC = 0.59), age (AUC = 0.63), and race (AUC = 0.57)—while maintaining moderate utility in predicting clinical endpoints. Further enhancement was achieved with the PP-VAE+LVEF variant, which slightly improved the prediction of age, reduced LVEF, and 5-year mortality, compared to the original PP-VAE, while still significantly curbing demographic information leakage. These findings underscore the potential of targeted representation learning to retain clinically relevant signal fidelity while reducing the model’s susceptibility to demographic attribute inference.

## Data availability

The data used in this study is part of an institutional research database with access restricted to institutional review board-approved researchers. The data cannot be shared publicly or made available to researchers at other institutions without a data use agreement. All of the codes generated in this study are available at https://github.com/PARC-KUMC/AI-Enabled-Privacy-Preserving-Cardiac-Diagnostics-via-Electrocardiograms

## Acknowledgements

The authors gratefully acknowledge the University of Kansas and the University of Kansas Medical Center for their support and facilitation of this study.

## Author contributions statement

A.N. and S.S participated in the conception and design of the study. FS.S. and C.H. conceived and conducted the experiment(s), FS.S., C.H., A.G., A.N., and S.S. analyzed the results. All authors reviewed the manuscript.

## Funding

This work was supported by the American Heart Association [Grant Number: 24TPA1291852]

## Competing interests

The authors declare no competing interests.

## Notes

### Competing Interest Statement

The authors have declared no competing interest.

### Funding Statement

This study was funded by American Heart Association

### Author Declarations

The study was approved as an expedited review by the Institutional Review Board at the University of Kansas Medical Center (STUDY00160252).

